# GenAI Exceeds Clinical Experts in Predicting Acute Kidney Injury following Paediatric Cardiopulmonary Bypass^2^

**DOI:** 10.1101/2024.05.14.24307372

**Authors:** Mansour Sharabiani, Alireza Mahani, Alex Bottle, Yadav Srinivasan, Richard Issitt, Serban Stoica

## Abstract

The emergence of large language models (LLMs) offers new opportunities to leverage, often unused, information in clinical text. This study examines the utility of text embeddings generated by LLMs in predicting postoperative acute kidney injury (AKI) in paediatric cardiopulmonary bypass (CPB) patients using electronic health record (EHR) text, and to explore methods for explaining their output. AKI is a significant complication in paediatric CPB and its prediction can significantly improve patient outcomes by enabling timely interventions. We evaluate various text embedding algorithms such as Doc2Vec, top-performing sentence transformers on Hugging Face, and commercial LLMs from Google and OpenAI. We benchmark the out-of-sample predictive performance of these ‘AI models’ against a ‘baseline model’ as well as an established clinically-defined ‘expert model’. The baseline model includes patient gender, age, height, body mass index and length of operation. The majority of AI models surpass, not only the baseline model, but also the expert model. An ensemble of AI and clinical-expert models improves discriminative performance by nearly 23% compared to the baseline model. Consistency of patient clusters formed from AI-generated embeddings with clinical-expert clusters - measured via the adjusted rand index and adjusted mutual information metrics - illustrates their medical validity. We use text-generating LLMs to explain the output of embedding LLMs, e.g., by summarising the differences between AI and expert clusters, and/or by providing descriptive labels for the AI clusters. Such ‘explainability’ can increase medical practitioners’ trust in the AI applications, and help generate new hypotheses, e.g., by correlating cluster memberships with outcomes of interest.

**Highlights:**

- LLMs outperform clinical experts in predicting risk of AKI after paediatric CPB.
- LLMs generate clinically plausible explanations and hypotheses using embeddings.
- Successful application of LLMs in paediatric CPB suggests potential in other specialised fields.
- Fine-tuning LLMs on domain data and forming ensembles of AI and clinical experts may boost accuracy.

## 1. Introduction

While traditional predictive models rely on structured data and/or expert-defined features, the emergence of large language models (LLMs) offers new opportunities to leverage unstructured text - e.g., clinical notes in electronic health records (EHRs) - for predictive modelling. Postoperative acute kidney injury (AKI) is a critical complication in paediatric patients undergoing cardiopulmonary bypass (CPB). Accurate prediction of AKI can significantly improve patient outcomes by enabling timely interventions. This study aims to assess the utility of text embeddings generated by LLMs in predicting AKI in paediatric CPB patients, and to explore methods for explaining their output and improving their predictive power.

Text embedding involves converting text — whether a single word, or a sequence of words forming a sentence, paragraph, or an entire article — into a numeric vector. Embedding techniques range from simple (e.g., bag-of-words, TF-IDF) to advanced (e.g., transformer-based LLMs). Modern text embedding models often produce high-dimensional vectors that are designed to preserve contextualised semantics of the input (natural language). Embedding a text string produces a vector, and thus embedding a text column results in a numeric matrix (Figure 1). Each column of this matrix can be treated as a feature in a predictive model.

**Figure 1:**
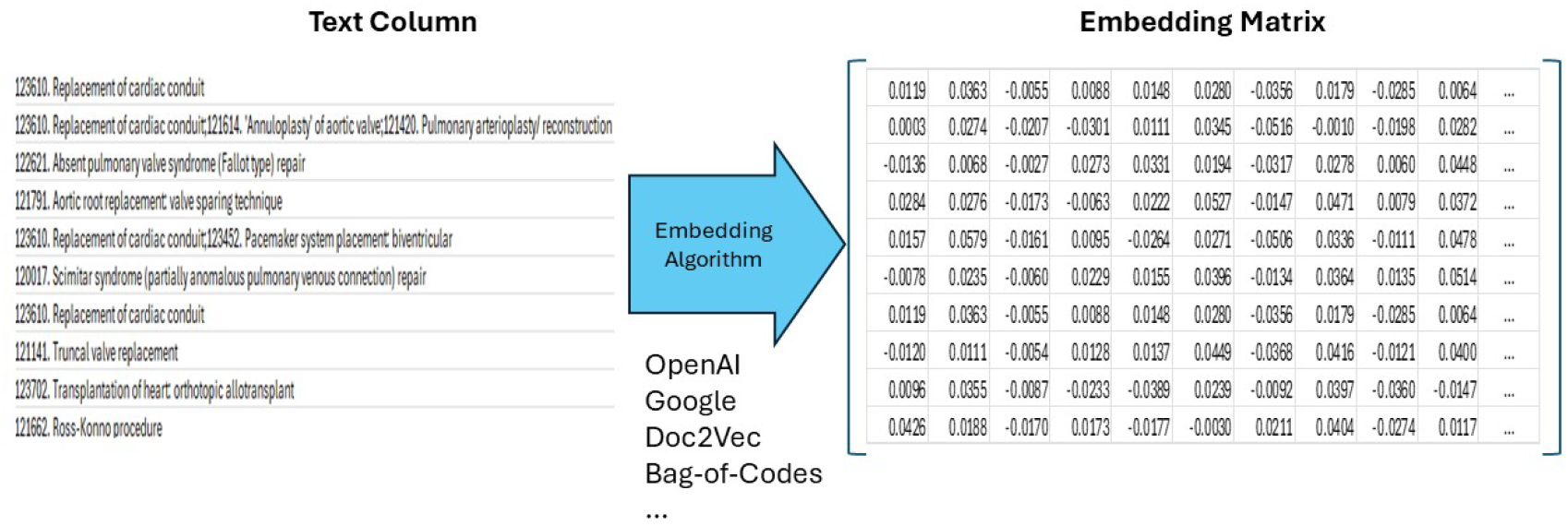
Illustration of the text embedding concept. Left: A text column representing the planned CPB procedures for 10 paediatric patients. Each entry is an ordered collection of standardised medical codes (numeric codes, followed by text). Right: The corresponding embedding matrix produced by OpenAI’s `text-embedding-3-large` model, with each observation represented by a high-dimensional, numeric vector (rows). While only 10 columns of the embedding matrix are shown here, modern LLMs produce vectors of much larger dimensions.

### 1.1. Related Work and Our Contributions

While text embeddings are often integral components of search engines, chatbots, information retrieval and recommender systems, their application in predictive models has received less attention. Pham et al [1] used embeddings derived from clinical text data to model patient trajectories and predict disease progression in individuals with diabetes and mental health conditions. Yan et al [2] conduct a systematic review of studies using unstructured clinical text in machine learning (ML) for prediction, early detection, and identification of sepsis. They conclude that combining text and structured data predicts sepsis earlier and more accurately than structured data alone. Kumar et al [3] use features extracted from word embeddings of clinical notes in classical machine learning as well as deep learning classifiers for predicting patient comorbidities. Khattak et al [4] provide a survey of word embeddings for clinical text, and discuss applications such as predicting unplanned readmission after discharge [5] and ICD code prediction [6].

While previous work has provided some evidence for the value of text embeddings in predictive tasks in medicine, our research makes several novel contributions. Firstly, we have included the latest public models from OpenAI and Google in our benchmarks, as well as several high-ranking sentence transformer models on Hugging Face, according to the Massive Text Embedding Benchmark (MTEB). This is important given the rapid pace of progress in the field. Secondly, our choice of problem and data (see Material and Methods) allowed us to use clusters defined by clinical experts as benchmarks for both intrinsic and extrinsic evaluation of clusters formed from LLM-produced text embeddings. Also, since each value in the text fields consists of one or more standardised medical codes (see, e.g., left side of Figure 1), we are able to apply a ‘Bag-of-Codes’ (BoC) encoding algorithm to each text field (see Material and Methods). By comparing the performance of BoC against LLM embeddings, we are able to assess the contribution of semantics and contextual information captured by the LLMs. Finally, our intuitive approach for leveraging text-generating LLMs to interpret the output of embedding LLMs is, to our knowledge, a practical ’explainability’ solution.

## 2. Material and Methods

### 2.1. Data Collection and Preparation

The data is based on a retrospective analysis of patients aged 18 and younger (maximum age treated at the institution), who underwent cardiac surgery with CPB at Great Ormond Street Hospital for Children, London. The cohort was selected using a National Institute for Cardiovascular Outcomes Research (NICOR) validated data set for patients operated on between April 2019 and April 2021. For further details on data collection, see [7].

For outcome, we focus on postoperative AKI. In particular, we consider the following five outcomes:

- *Creatinine ratio*: Ratio of postoperative to baseline serum creatinine.
- *Urine output (0*.*5)*: Length of time (in hours) within the 48 hours after CPB when patient’s urine output per hour is below 0.5 ml per kilogram of body mass.
- *Urine output (0*.*3)*: Same as above, but comparing urine output to a 0.3 ml per kilogram of body mass (per hour) threshold.
- *KDIGO-Ordinal* : An integer, ordinal score from 0 to 3, defined from using a combination of the above three numbers, per KDIGO protocol [8].
- *KDIGO-Binary*: A binary indicator of severe AKI. It is 1 if KDIGO-Ordinal is 2 or more, and 0 otherwise.

(For predictive tests, we focused on ‘KDIGO-Binary’ as outcome, while for explainability analyses, we considered all five outcomes.)

A set of ‘baseline’ patient attributes were included in this analysis: gender, age, height and weight, as well as the length of operation. Note that age, height and weight were all recorded as of the start of operation. Weight and height were transformed to BMI and ‘residual’ height. The former followed the standard definition, while the latter was defined as the percent deviation of height from the predictions of a LOESS (locally estimated scatterplot smoothing) model that regressed height on age and gender.

The EHR text data included patient diagnoses (`diagnosis`), and operations performed (`operation`). Each text field value is an ordered set of standardised medical codes, with first entries representing primary codes. Expert-defined clusters for `diagnosis` (called, ‘broad diagnosis grouping’ or BDG) and `operation` (called, ‘transformed specific procedure’ or TSP) were also obtained using the Partial Risk Adjustment in Surgery (PRAiS) v2 protocol [9]. PRAiS is a model developed in the UK to predict 30-day mortality risk after paediatric heart surgery. The BDG and TSP groupings are defined and used in the PRAiS risk model as categorical variables.

After removing operations with missing values, we are left with 780 data points. Table 1 provides a summary of the data.

**Table 1:** Summary of dataset used in this paper. Number is parentheses indicate percentages of total data. Numbers in brackets represent the interquartile range. Note that for 30-day mortaility calculation, sample size is slightly smaller.

| **Variable** | **Value** |
|---|---|
| Number of operations | 780 |
| Number of unique patients | 736 |
| KDIGO-Ordinal |  |
| 0 | 315 (40) |
| 1 | 266 (34) |
| 2 | 121 (16) |
| 3 | 78 (10) |
| KDIGO-Binary |  |
| 0 | 581 (74) |
| 1 | 199 (26) |
| Median age (months) at operation | 8.3 [3.1 - 43.5] |
| Range | 0.1 - 188.3 |
| Median BMI at operation | 15.3 [13.6 - 16.7] |
| Range | 8.3 - 32.9 |
| Mean/std of height residual | -0.001/0.07 |
| Median operation time (minutes) | 86 [57 - 122] |
| Range | 10 - 471 |
| 30-day mortality (757 operations) |  |
| Observed | 2 (0.26) |
| Predicted (PRAiS) | 11 (1.5) |

### 2.2. Software and Compute Environment

The initial data preparation steps were performed in R 4.4.0 [10]. All remaining steps were performed in Python 3.11.7. All Python and R scripts were executed on a 64-bit Windows 11 PC running on Intel(R) Core(TM) 7 150U 1.80 GHz, with 16 GB of installed RAM.

### 2.3. Text Embeddings

Each text entry consisted of one or more standardised medical codes. Each medical code entry is an integer, followed by a text description. The order of the codes has medical significance: The most important codes - e.g., patient’s primary diagnosis - are listed first. An example of an entry for the `diagnosis` field is: `155516. Cardiac conduit failure;103604. Aortic regurgitation - acquired;090101. Common arterial trunk`.

Each text field entry is transformed into a numeric vector, using various encoding/embedding algorithms:

#### 1. Bag-of-Codes (BoC)

Each medical code gets a binary indicator, and each text field entry gets 1’s for all medical codes present in the string, and 0’s for others. The output dimension is thus dictated by the number of distinct medical codes used for each field, which is 282 and 179 for `diagnosis` and `operation`, respectively. This is a generalisation of one-hot encoding, where multiple levels of a categorical variable are not mutually exclusive. This is also similar to the standard bag-of-words (BoW) approach, where we treat each medical code as a word. (However, unlike BOW, in BoC the entries are binary, i.e., the same code cannot appear more than once in a text field.) Note that using the BoC method is possible in our problem since - as illustrated earlier - the text fields are not free-format, but rather a collection of standardised entries. BoC serves as a reference, against which more sophisticated embedding algorithms (discussed below) are benchmarked.

#### 2. Doc2Vec [11]

We used the implementation by the `gensim` Python package. Both distributed bag-of-words (DBOW) and distributed memory (DM) versions were tested, using values of 10, 100, 1000 for number of training epochs, and values of 40, 400 and 4000 for output dimension (i.e., length of embedding vector).

#### 3. Open-Source LLMs

We used the publicly-available, top-ranking models from the MTEB leaderboard for embedding models [12], plus `pubmedbert-base-embeddings-matryoshka` (which is a medically-specialised embedding model), `all-MiniLM-L6-v2` (which is a small model built on a distilled, pre-trained model) and `ClinicalBERT`. The models were downloaded from the Hugging Face repository using the `sentence-transformers` Python package. With the exception of the last model, all others have been fine-tuned for embedding tasks. For `ClinicalBERT`, we added mean pooling to the last hidden layer to convert token embeddings to sentence embeddings. While it is technically more appropriate to refer to the sentence-transformer model built on `ClinicalBERT` as `ClinicalBERT+mean pooling`, hereafter we will refer to this model as simply `ClinicalBERT` for brevity.

#### 4. Commercial LLMs

We used OpenAI and Google embedding LLMs via their web APIs, accessed through Python packages `openai` and `vertexai`, respectively. OpenAI offers three embedding models, `text-embedding-3-large`, `text-embedding-3-small` and `text-embedding-ada-002`, with output dimensions of 3072, 1536 and 1536, respectively. Hereafter, and for brevity, we will refer to them as `OpenAI-large`, `OpenAI-small` and `OpenAI-ada`, respectively. For Google, we used two models, `text-embedding-004` (or `Google-004` for short), and textembedding-gecko@003 (`Google-gecko` for short), both with an output dimension of 768. To use Google’s embedding models, we must also specify a ‘task’ parameter, with options being `RETRIEVAL QUERY`, `RETRIEVAL DOCUMENT`, SEMANTIC SIMILARITY, `CLASSIFICATION`, `CLUSTERING`, `QUESTION ANSWERING` and `FACT VERIFICATION`. (The last three tasks are only applicable to the `Google-004` model.)

## 2.4. Fine-Tuning

We curated 1593 title/abstract pairs from the PubMed online database, using the keywords ‘acute kidney injury AND cardiopulmonary bypass’. (PubMed was accessed using the `Bio` Python package.) These pairs were used to fine-tune three LLMs: `ClinicalBERT`, `PubMedBERT-embeddings` and `MiniLM-L6-v2`. The loss function used was `MultipleNegativesRankingLoss` in the `sentence-transformers` library.

In a nutshell, this loss function forces the embedding vectors for matching title/abstract pairs to be more similar (measured by their cosine distance) than non-matching pairs formed by random shuffling of titles and abstracts in the same batch. We used the following parameters for fine-tuning the LLM: maximum number of epochs (20), learning rate (1e-6), warmup ratio (0.1), fraction of data used for early stopping (0.25), early-stopping patience (3 epochs), early-stopping threshold (0.0).

## 2.5. Spherical K-Means Clustering

We use ‘spherical’ k-means, which we implemented in Python, for clustering patients according to their text embedding vectors. This is a variant of standard k-means, where the metric used to measure the distance between two vectors **x** and **y** is changed from L2 norm (i.e., (**x** − **y**)^*T*^ (**x** − **y**)) to ‘cosine distance’, *D*_*C*_(**x, y**):

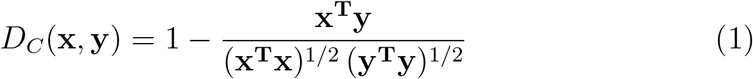

This metric is invariant with respect to the length of the two vectors, focusing only on their angles and thus making it a suitable choice for text embedding vectors that are L2-normalised, i.e., **x**^**T**^**x** = 1. (Note that while L2 and cosine distances would produce identical assignments of points to clusters, they would differ in the centroid update step of the k-means clustering algorithm.) One exception is the BoC method, which does not produce L2-normalised vectors (in fact the number of medical codes present in the text field for a given patient - which is equal to the L1 norm of the BoC vector - has potentially relevant information), and hence we applied standard k-means clustering to BoC embeddings, rather than spherical k-means.

## 2.6. Cluster Similarity Metrics

We use two symmetric metrics to quantify the degree of similarity between two clustering algorithms: 1) adjusted rand index (ARI) [13], and 2) adjusted mutual information (AMI) [14]. In both cases, the adjustment subtracts a baseline value to account for the possibility of ARI/AMI values occurring by chance, and also divides by the possible range of the value to achieve a normalised score that has an upper bound of 1, and produces values close to 0 for two clusters with no concordance. Calculations were done using the `metrics` module of the `scikit-learn` Python package.

## 2.7. Predictive Contribution of Embeddings

To quantify the contribution of text embeddings towards predicting AKI, we perform spherical k-means clustering - as discussed earlier - on the embedding vectors produced by the LLMs and other embedding algorithms listed in Text Embeddings. We then proceed to include the cluster ‘soft’ membership data as features - alongside the baseline features mentioned in Data Collection and Preparation - in a binary classification model. We compare the area under ROC (AUC) of models trained on cluster features based on different embedding models (the ‘AI models’) against each other, as well as against the model using expert clusters instead of embedding-based clusters (the ‘expert model’). In both types of models, we separately train two models, one using the `diagnosis` clusters alongside the baseline variables, and one using the `operation` clusters added to baseline variables. The predicted probabilities of these two models are averaged to form the final prediction.

## 2.8. Hyperparameters

As mentioned in Text Embeddings, two of the embedding algorithms that we used have hyperparameters: Doc2Vec (number of training epochs, output dimension) and Google (embedding task). In both cases, we calculate the average AUC across all CV folds for each hyperparameter value or combination (and within each sub-model, such as the DBOW and DM versions of Doc2Vec), and choose the hyperparameter value that produced the highest AUC.

There are two other important hyperparameters in our experiments. First is the number of clusters in (spherical) k-means. For cluster consistency expeirments, we chose this to be equal the number of expert clusters (which is 11 for `diagnosis` and 15 for `operation`). This removes the confounding effect of the number of clusters from calculations of the ARI/AMI consistency metrics. In the predictive models, we used a round value of 10 for both text fields. The second hyperparameter is the number of dimensions of the embedding vector to include in downstream steps. For instance, one can choose the first 100 elements of a 768-dimensional embedding vector, or apply dimensionality-reduction techniques such as Principal Components Analysis. Here, we chose to always feed the entire embedding vector to the downstream clustering algorithms. Tuning these two hyperparameters and studying their effect on consistency of clusters with experts and/or predictive performance of models will be a topic of future research.

## 2.9. Ensemble Models

We used a simple, weighted-average ensemble approach, applied to the predicted probability of severe AKI produced by each constituent of the ensemble model. In the ‘AI Ensemble’, two models were included with 2:1 weights: OpenAI’s `text-embedding-3-large` (weight of 2) and Google’s textembedding-gecko@003 using the task `CLASSIFICATION` (weight of 1). In the ‘AI + Expert’ ensemble, the Expert model was added to the AI ensemble, with a weight equal to 1/10th of OpenAI’s weight. These weights were chosen based on limited trial-and-error. Applying more sophisticated ensemble techniques is another topic for future research.

## 2.10. Explainability

Text-generating LLMs were used in two explainability roles. First, we asked OpenAI’s `gpt-4o` model to summarise the differences between AI clusters that were created by applying spherical k-means to the `operation` embeddings, and the corresponding expert clusters (TSP). A detailed description of the prompt and LLM response is provided in Appendix A.

Secondly, we asked the same OpenAI model as well as Google’s `gemini-1.5-pro` model to provide descriptive and distinct labels for clusters formed from a concatenation of `diagnosis` and `operation` embeddings. In this task, we used the ‘structured output’ features of OpenAI and Google to ensure that the LLM response was formatted in a predictable structure. This allows for automated creation of downstream tables and figures. Details can be found in Appendix B. This explainability task is completed by computing the rank correlation - using Kendall’s tau - of the five AKI-related outcome metrics defined in Section 2.1 vs. membership in each of the embedding-based clusters.

Note that both of the above explainability tasks involve long prompts (e.g. *>* 50,000 tokens) that would exceed all but the latest generations of OpenAI and Google models which are equipped with long context windows.

## 2.11. Benchmarking

For quantifying the consistency of AI and expert clusters, we took the average ARI/AMI scores from 10 runs of spherical k-means. Within each run, we used 10 random initialisations of cluster centroids, where the cluster with the smallest total within-cluster distance is selected. Number of clusters was pre-selected to match the number of expert clusters (11 for `diagnosis` and 15 for `operation`).

In predictive experiments, we used 250-times repeated, 10-fold cross-validation to compare the AUC of different models. To maximise comparability of results, we fixed the random seed when calling the `RepeatedKFold` function of `scikit-learn`. For each run, therefore, we obtained 2500 AUC numbers, which we use to perform statistical analyses such as paired t-test.

To quantify the stability of AI clusters, we applied spherical k-means to concatenated embeddings of `diagnosis` and `operation` fields, produced by the `OpenAI-large` model. This was repeated 10 times, each time using 100 random starts and 10 clusters. We then calculated average ARI/AMI scores between all 45 (10*×*9/2) unique pairs of clustering runs. The clusters were mapped across runs using a greedy algorithm for maximising member overlap. See Appendix C for the source code and explanation of the cluster mapping algorithm.

Next, for each of the 10 clustering runs, we used a similar process to that outlined in the Explainability to solicit cluster labels from OpenAI and Google models. We split the 10 clustering runs evenly between OpenAI and Google. Labels were aligned across runs using the mapping generated per above.

## 3. Results

### 3.1. Consistency of AI and Expert Clusters

Table 2 shows the ARI and AMI scores between AI clusters formed from `operation` and `diagnosis` text fields and their expert counterparts. The positive values of these metrics - which remove a baseline effect due to chance - indicate a statistically significant, although imperfect, consistency between the two approaches in extracting information from the underlying text fields. Further evidence for sensibility of AI clusters is that AI and expert clusters corresponding to the same text field (`operation` or `diagnosis`) are more consistent than clusters based on non-matching fields. At the same time, the fact that non-matching clusters also have non-zero consistency can be explained by the correlation between a patient’s diagnoses and the operations performed on them.

**Table 2:** Consistency between ‘expert’ clusters (rows) and ‘AI’ clusters (columns), measured by the ARI and AMI metrics (1st/2nd numbers in each cell, respectively). AI clusters are generated by applying spherical k-means to text embedding vectors produced by OpenAI-large. Expert clusters are defined by the PRAiS v2 risk model. See Material and Methods for further details.

|  | **AI Clusters** |  |
|---|---|---|
| **Expert Clusters** | Diagnosis | Operation |
| Diagnosis (BDG) | **0.31 / 0.44** | 0.21 / 0.32 |
| Operation (TSP) | 0.27 / 0.40 | **0.29 / 0.48** |

Figure 2 shows the average ARI and AMI scores for `diagnosis` and `operation` fields using embeddings produced by different LLMs. The mean and 95% confidence intervals for the BoC approach are included as horizontal lines for comparison. We see that the majority of LLMs produce clusters that have higher consistency with expert clusters, compared with BoC.

**Figure 2:**
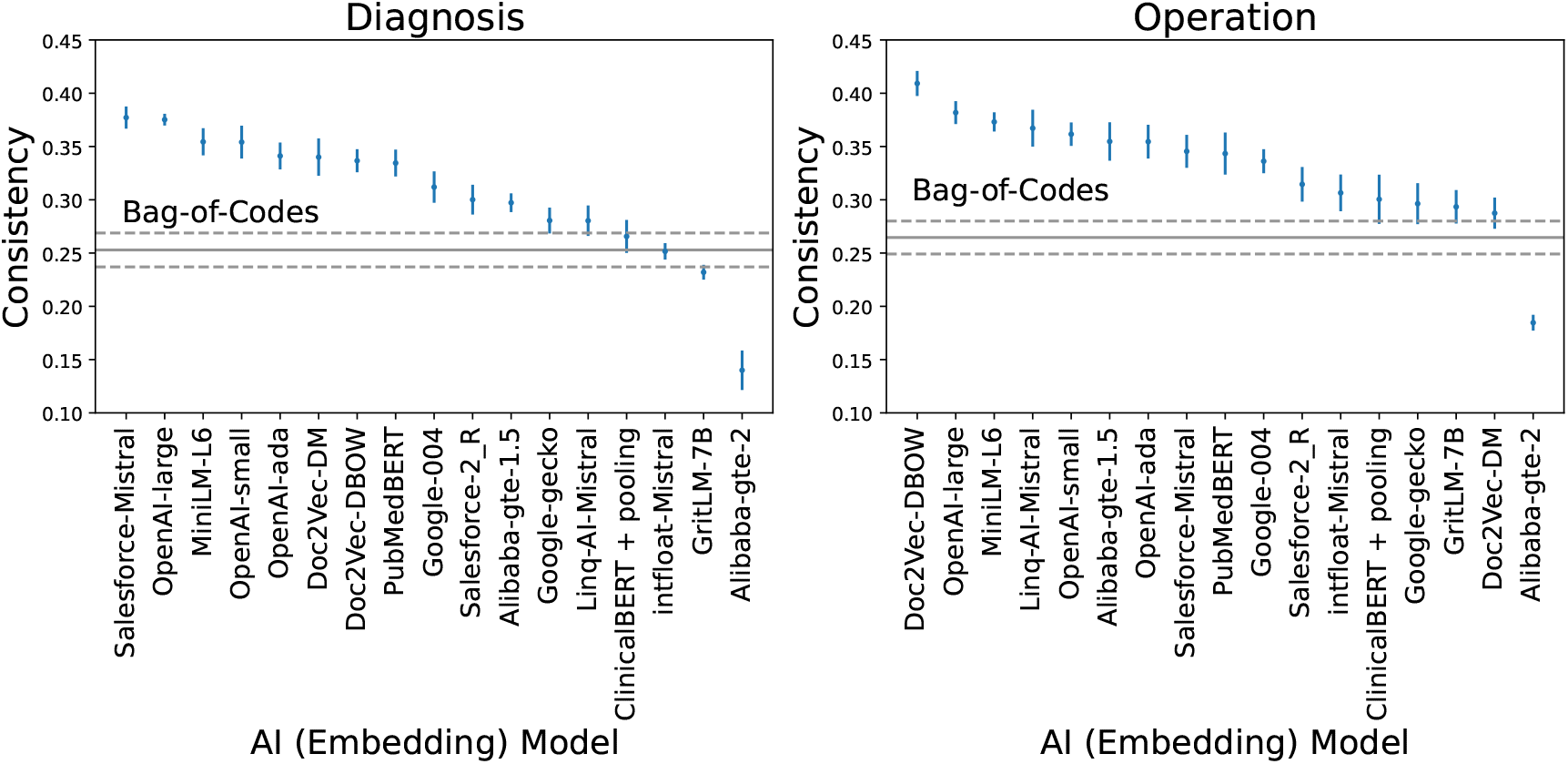
Consistency of AI and expert clusters based on `diagnosis` (left) and `operation` (right) text fields. Consistency is defined as the average of ARI and AMI scores between AI and expert data partitions. Confidence intervals are based on the 10 repeats of k-means clustering, each using 10 random initialisations of centroids. For Bag-of-Codes (BoC), we used standard k-means, while spherical k-means was used for AI embeddings. Horizontal lines represent mean (solid line) and 95% confidence intervals (dashed lines) for the BoC method. Error bars for each embedding model also represent the 95% CI.

`OpenAI-large` has the highest combined ranking across both text columns, i.e., most consistency with experts. Since BoC - unlike domain experts - is not context aware and does not use the information captured in the order of entries of multiple codes for a single patient, our results suggest that LLMs are successful at extracting medically-relevant information using the context provided by the entire text string, including the order of code entries.

We asked OpenAI’s `gpt-4-turbo` model to summarise and explain the differences between AI and expert clusters. As seen in the excerpt below, it produced an insightful and plausible answer:

*Partition 2 [experts] appears more segmented, potentially dividing patients by broader surgical categories, like major surgery types (Aortic arch repairs, Tetralogy of Fallot repairs, Norwood procedures, etc*.*). This suggests a more high-level clustering compared to the specific procedural focus in Partition 1 [AI]* …

Please note that the partition labels in square brackets are added by us; in other words, we did not relay any information to the LLM about which data partition corresponds to AI and which one corresponds to experts. Appendix A contains the full text of the prompt provided to the LLM and its response.

### 3.2. Predictive Performance of Embeddings

Figure 3 compares the area under receiver operating characteristic curve (AUC) for predicting ‘AKI-Binary’ using various embedding approaches (as well as the ‘No Embedding’ and ‘Ensemble’ options) against the ‘Expert Model’. Among the individual algorithms tested, `OpenAI-large` (second bar from left) outperforms all others, including the expert model (horizontal line). The `ClinicalBERT` model, despite not being fine-tuned for sentence embedding, shows competitive performance, being the top open-source model and only behind OpenAI and Google.

**Figure 3:**
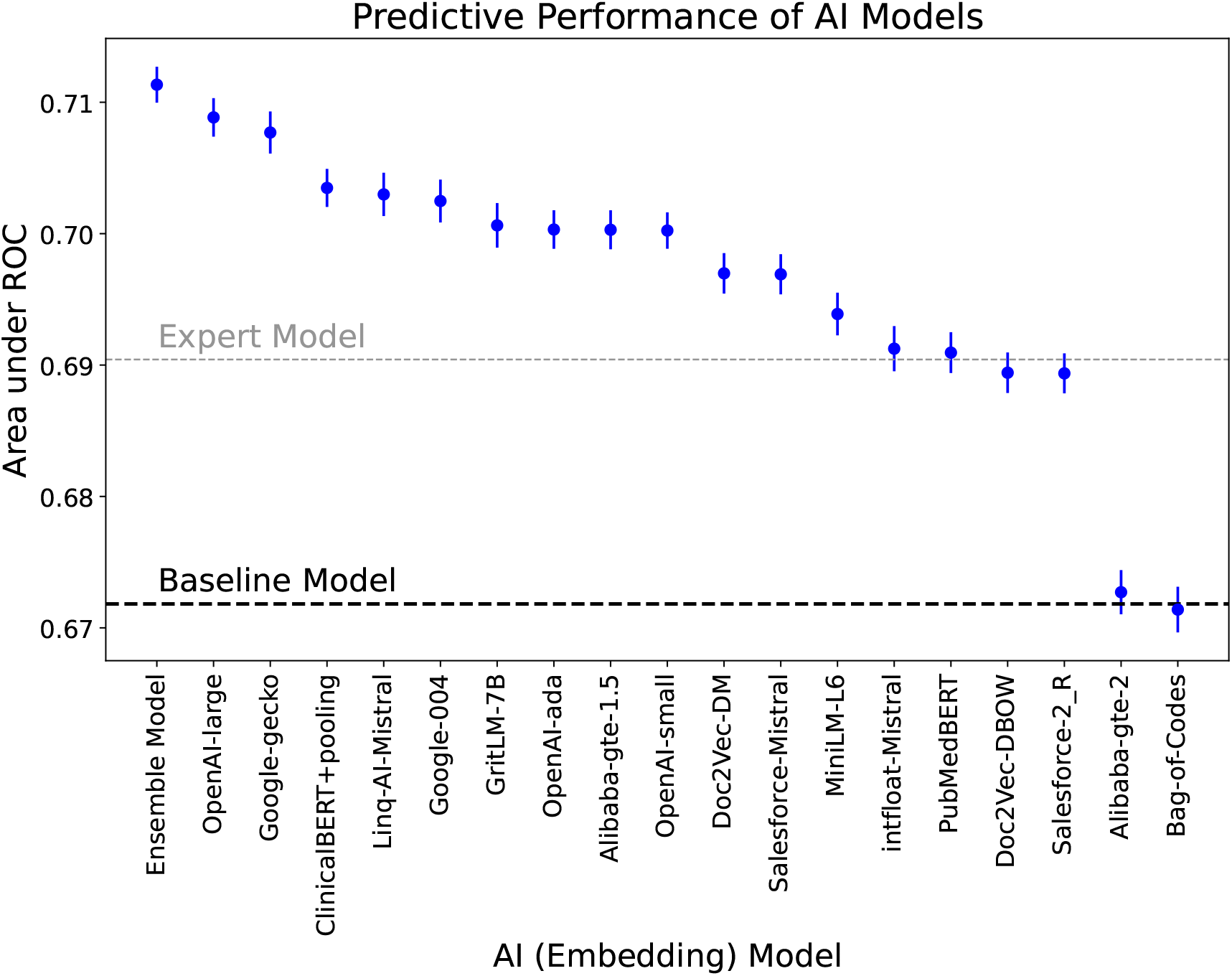
Out-of-sample AUC of binary classifiers using clusters generated - via spherical k-means - from text embedding vectors produced by various LLMs. The horizontal lines represents the performance of a baseline model that does not use any embeddings, as well as an expert-based model, which takes the average predicted probability of two binary classifiers, each using expert clusters for diagnosis and operation as categorical variables, alongside baseline attributes. ‘Ensemble Model’ predictions are a weighted average of ‘OpenAI-large’, ‘Google-gecko’ and ‘Expert’ models.

Forming an ensemble of AI and expert models further improves performance (leftmost bar). This is expected since, while AI models exhibit high correlation of fold-level errors amongst themselves, their correlation with the expert model is significantly smaller. For instance, average correlation between pairs of AI models in the group is 92.8%, while their average correlation with the expert model is 83.0%. This is in line with results presented earlier in Table 2 showing an imperfect consistency between the AI and expert clusters.

Compared to the baseline model, we achieved nearly 23% discriminative power improvement corresponding to AUC improvement from 67.2% (baseline model) to 71.1% (the final model).

We also tested the impact of fine-tuning three open-source LLMs on the predictive performance of models trained on their text embeddings. Results are shown in Table 3. For two of the three models tested, there is modest performance improvement, while for the third model - which has the highest pre-tuning performance - we see performance degradation.

**Table 3:** Impact of fine-tuning on AUC (in percentage terms) of models trained on clusters formed - via spherical k-means - from three open-source embedding models. P-values are based on paired t-test of fold-level errors, formed from 250-times repeated, 10-fold cross-validation.

| **LLM** | **Before** | **After** | **Change** | **P-value** |
|---|---|---|---|---|
| PubMedBERT-embeddings | 69.09 | 69.20 | 0.11 | 1e-3 |
| MiniLM-L6-v2 | 69.39 | 69.54 | 0.15 | 2.8e-5 |
| ClinicalBERT | 70.35 | 69.85 | -0.50 | < 1e-5 |

### 3.3. AI Explainability

Average ARI/AMI scores across all 45 pairs of 10 clustering runs described in Benchmarking are 83% and 89%, respectively, indicating high stability of AI-generated clusters across runs. Similarly, after mapping the clustering runs using the algorithm described in Appendix C, the average member overlap across all pairs is 86%.

Cluster labels produced by OpenAI and Google - as well as their oddsratios - were quite consistent. Table 4 shows cluster labels for two OpenAI and two Google runs. The full mapping table is provided as a supplementary material (see Appendix D).

**Table 4:** Comparison of two cluster labels produced by OpenAI and two by Google, each produced for clusters that were outpu1t9of spherical k-means with a different set of 100 random initialisations. Cluster numbers across runs 1-3 were mapped to run 0 to maximise their member overlap using a greedy algorithm described in the Appendix. The group highlighted in red shows statistically significant association with increased AKI risk, while the group highlighted in green shows significant association with reduced AKI risk. ‘Short Label’ column matches x axis values in Figure 3.

| Short Label | OpenAI 1 | Google 1 | OpenAI 2 | Google 2 |
|---|---|---|---|---|
| Conduit Issues and Repairs | Cardiac Conduit Issues and Associated Repairs | Valve replacement or repair | Cardiac Conduit Failure and Replacement | Valve Replacement & Repair |
| Pulmonary Atresia & Fontan | Complex Pulmonary Atresia and Fontan Procedures | Univentricular heart or HLHS procedures | Complex Congenital Heart Disease | Single Ventricle Physiology |
| ASD Repairs | Atrial Septal Defect (ASD) Repairs | ASD closure | Atrial Septal Defect (ASD) Management | Atrial Septal Defects |
| VSD Repairs | Ventricular Septal Defect (VSD) Repairs | VSD closure | Ventricular Septal Defect (VSD) Treatment | Ventricular Septal Defects |
| Outflow Tract & Valve | Complex Outflow Tract Obstructions and Valve Replacements | Complex congenital heart defect repair | Anomalous Pulmonary Venous Connections | Complex Congenital Heart Defects |
| TGA Surgeries | Transposition of Great Arteries and Associated Surgeries | Arterial switch for TGA | Transposition of the Great Arteries (TGA) Management | Transposition of the Great Arteries |
| Aortic Valve Repairs | Aortic Valve Stenosis and Regurgitation Repairs | Aortic valve or arch repair | Aortic Valve and Outflow Pathway Surgery | Aortic Valve Abnormalities |
| AVSD & Related Repairs | Atrioventricular Septal Defect (AVSD) and Related Repairs | AVSD repair | Atrioventricular Septal Defect (AVSD) | Atrioventricular Septal Defects |
| Heart Transplants | Heart Transplant and Advanced Cardiac Therapies | Transplant, assist device, or complications | Heart Transplant and Complications | Heart & Lung Transplant / VAD |
| Fallot & Related Repairs | Tetralogy of Fallot and Related Repairs | Tetralogy of Fallot repair | Tetralogy of Fallot and Associated Repairs | Tetralogy of Fallot |

We can gain further insight by considering how the underlying metrics that feed into the calculation of the KDIGO score correlate with the AI-generated clusters. Figure 4 summarises this analysis. We make a few observations:

**Figure 4:**
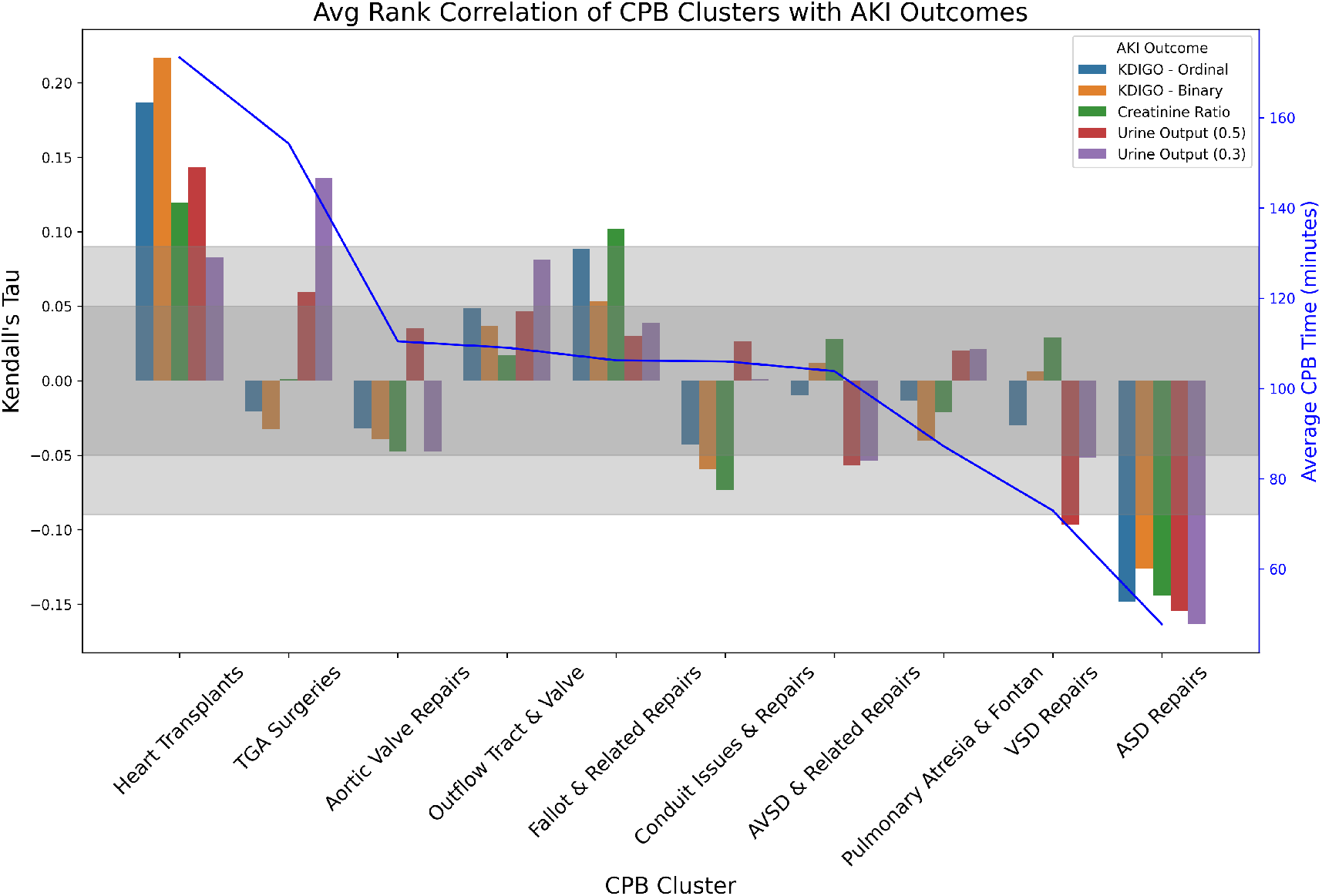
Average rank correlation (measured via Kendall’s tau) of AKI-related outcomes with membership in each of the 10 clusters defined by applying spherical k-means to a concatenation of `OpenAI-large` model’s embedding of `diagnosis` and `operation` text columns. Each clustering run is subsequently labelled 10 times by text-completion LLMs, five times using OpenAI’s `gpt4-turbo` model and five times using Google’s `gemini-1.5-pro` model. See Section 2.1 for definition of AKI metrics. Clusters are sorted from left to right in decreasing average operation time, which is displayed as a blue line (right y axis). Inner grey band reflects the 95% confidence interval, outside of which correlations are statistically significant. The outer grey band is similar, but after Bonferroni correction for multiple testing, using 50 tests (5 outcome variables *×* 10 clusters). Note that cluster (x axis) labels are abbreviated versions of the longer, more descriptive cluster labels produced by the LLMs, as listed in Table 4.

- Two groups show consistent, statistically significant deviation from the rest: heart transplants show higher risk for all AKI outcomes vs. other CPB clusters, and ASD repairs show lower risk than the rest.
- For TGA, there is a higher risk of low urine output (especially for length of time below 0.3 ml/kg/hr) that is statistically-significant, even after adjusting for multiple testing. However, this is not reflected in KDIGO outcomes and their changes are statistically insignificant.
- For the Fallot group, serum creatinine ratio is significantly higher than the rest, and ordinal KDIGO score is also nearly significant (after multiple-testing adjustment).
- For the Conduit group, we see protection in terms of creatinine ratio, which is borderline significant.
- For the VSD group, we see significant protection in terms of urine output.

All of the above observations fit with intuition of clinical experts (co-authors of this paper) and are also consistent with what is being observed in practice.

## 4. Discussion

This study demonstrates the value of LLMs in predicting postoperative AKI in paediatric CPB patients. The superior performance of LLMs, even in a highly specialised domain, underscores their versatility and power. Prediction may also be further improved by forming ensembles of multiple LLMs and clinical expert models. On the other hand, we show that text-generating LLMs can successfully summarise the output of embedding LLMs, resulting in explainable AI classifications. This is a promising way to enhance trust in the use of AI prediction in medical settings and to provide deeper insights into the clinical questions.

In our experiments with embedding vectors, we used their full length in downstream clustering algorithms. Alternatively, we could explore the effect of applying dimensionality-reduction to embedding vectors. With many recent embedding models having been trained using the Matryoshka loss function [15], a stand-alone dimensionality-reduction step may not even be necessary. Yet the number of dimensions to keep would still be a hyperparameter that requires tuning - e.g. via grid search - as part of the overall predictive modelling process.

Weighted-ensemble approach is effective; however, there is room for improvement. Firstly, we only included two AI models in the ensemble; adding more models to the ensemble could provide further improvement. Also, determining the weights can be done more systematically, e.g. using ensemble techniques such as stacked generalisation [16]. However, the main focus of this paper was not investigating ensemble of AI models.

Latest advances by two leading commercial providers of AI - OpenAI and Google - greatly facilitated our ‘explainability’ framework. Firstly, the increased context length of the LLMs (128k tokens for OpenAI’s `gpt-4o` and *>* 2 million tokens for Google’s `gemini-1.5-pro-001`) was key to our ability to execute the cluster labeling task in a single pass. This is because our prompt had in excess of 50k tokens, due to a large payload that included the text columns of interest for the entire training data. For LLMs with short context length, a potential solution would be to implement a multi-stage approach, e.g., by first creating labels for each cluster in isolation, and wrapped in a for loop, and then doing a second round of interaction with the LLM to tune the labels and make them more differentiated.

Secondly, we took advantage of the structured/JSON output modes offered by OpenAI and Google models. Enforcement of a data structure allows for automation of downstream tasks, which in our case amounted to creating summary tables showing rank correlation of membership on each cluster with outcomes. Such table summaries were the foundation for creation of Figure 3.

One potential limitation of our work is that we we were not able to use any text information that could possibly lead to patient identification. This limitation is likely to be repeated in practice given the utmost importance of respecting patient confidentiality. Most likely, adding further text data would have resulted in much better prediction and explainability and clinical insight. On the other hand, this limitation led us to use text fields that contain standardised medical codes - for patient diagnoses and procedures - which, in turn, allowed us to benchmark our prediction and clinical findings against existing clinical expert classification.

Our limited experiments with fine-tuning point to its potential role in improving predictive performance. While we indirectly incorporated the outcome variable by including the AKI keyword in our PubMed search, yet we did not use our training data - text or outcome columns - directly in the fine-tuning process. This and other variations on fine-tuning are the subject of future research.

While we have illustrated the value of Generative AI in predictive modelling, many practitioners may be reluctant to use LLMs due to perceived or real technical barriers. To help overcome such barriers, we will be releasing a Python implementation of various steps discussed in this paper, including embedding of a text column, applying spherical k-means, and soliciting cluster interpretation from LLMs.

## 5. Conclusion

Rapid advances in quality, accessibility and user-friendliness of modern LLMs indicate a promising future for their application in predictive analytics, including in specialised medical domains. Their ability to match or surpass clinical expert models, coupled with our proposed solutions for explainability and performance improvement, positions LLMs as valuable assets in clinical research and practice. Future work will focus on applying our lessons and techniques in other medical domains, and exploring additional strategies to further enhance their explainability and predictive performance.

## Supporting information

Cluster mappings - full results

## Data Availability

All data produced in the present study are available upon reasonable request to the authors.

## 7. CRediT authorship contribution statement

**Mansour Sharabiani**: Conceptualisation, Formal analysis, Investigation, Methodology, Software, Visualisation, Writing – original draft, Writing – review and editing; **Alireza Mahani**: Conceptualisation, Formal analysis, Investigation, Methodology, Software, Visualisation, Writing – original draft, Writing – review and editing; **Alex Bottle**: Supervision, Validation, Writing – review and editing; **Yadav Srinivasan**: Data curation, Validation, Writing - review and editing; **Richard Issitt**: Data curation, Validation, Writing - review and editing; **Serban Stoica**: Supervision, Validation, Writing – review and editing

## 8. Use of Generative AI in the Writing Process

During the preparation of this work the authors used ChatGPT for editing the manuscript. After using this tool, the authors reviewed and edited the content as needed and take full responsibility for the content of the published article.

## 10. Glossary

Acute Kidney Injury (AKI): A sudden decrease in kidney function, often occurring after surgery, particularly in paediatric patients undergoing cardiopulmonary bypass (CPB).
Adjusted Mutual Information (AMI): A measure of agreement between two clusterings, adjusted for chance, based on the mutual information between the clusterings.
Adjusted Rand Index (ARI): A metric used to measure the similarity between two data clusterings, adjusted for the chance grouping of elements.
Area Under the Receiver Operating Characteristic Curve (AUC): A performance measurement for classification models at various threshold settings, indicating the ability of the model to distinguish between classes.
Bag-of-Codes (BoC): A text embedding technique where each medical code in a patient’s record is represented as a binary indicator in a vector.
Cardiopulmonary Bypass (CPB): A technique used during heart surgery where a machine temporarily takes over the function of the heart and lungs, allowing surgeons to operate on a still heart.
Cross-Validation (CV): A statistical method used to estimate the performance of machine learning models, where the data is split into multiple folds, and the model is trained and validated on different folds.
Doc2Vec: A text embedding technique that learns distributed representations of documents, allowing for the transformation of entire documents into fixed-length vectors.
Ensemble Model: A machine learning technique that combines the predictions of multiple models to improve accuracy and robustness.
Explainability: Techniques used to interpret and understand the predictions made by complex machine learning models, often to increase trust and provide insights into the decision-making process.
Fine-Tuning: The process of adjusting a pre-trained model on a new dataset, typically with a smaller learning rate, to adapt the model to a specific task or domain.
Hyperparameters: Parameters of a machine learning model that are set before training and control the learning process, such as the number of clusters in k-means or the learning rate in neural networks.
KDIGO: Kidney Disease Improving Global Outcomes; a set of guidelines used to define and classify the severity of acute kidney injury.
Large Language Models (LLMs): Advanced machine learning models, often based on transformer architectures, that are trained on vast amounts of text data and can perform a variety of natural language processing tasks.
Partial Risk Adjustment in Surgery (PRAiS): A model used in the UK to predict 30-day mortality risk after paediatric heart surgery, incorporating various clinical variables.
Spherical K-Means: A variant of the k-means clustering algorithm that uses cosine distance instead of Euclidean distance, making it suitable for clustering high-dimensional data like text embeddings.
Text Embedding: A method of converting text into numeric vectors that capture the semantic meaning of the text, used in machine learning models for various predictive tasks.

## Appendix A. AI Explaining AI - Part 1

### Appendix A.1. Prompt

The following shows the prompt preamble and sample data that follows it, asking OpenAI’s `gpt-4-turbo` model to interpret the difference between two partitions of patients according to their planned operations. One partition was generated by applying spherical k-means to embeddings of the `operation` field by the `OpenAI-large` embedding model (‘AI cluster’). The other partition was created by using the PRAiS v2 protocol (‘expert cluster’). As can be seen below, the prompt did not reveal to the LLM which partition corresponded to AI/expert cluster:

> A group of pediatric patients have undergone cardiopulmonary bypass surgery (CPB). For each patient, we have one or more numeric codes and their associated descriptions concatenated via ’;’ into a single sting. Each patient is printed on a single line. The codes represent one or more surgical procedures performed during the CPB. The dataset has been partitioned into 15 groups by two different clustering methods (Partition 1 and Partition 2), based on the aforementioned text entries for the patients. Clusters of patients in each partitioning scheme are separated by multiple blank lines. The two partitioning of patients are printed below. Note that the input to the two partitioning schemes has been an identical set of patients, with each patient having the same exact text entry in both cases. Please provide a brief interpretation of the most salient differences between the two partitions:
>
> Partition 1:
>
> 120102. Atrial septal defect (ASD) secundum closure with direct suture
>
> 120103. Atrial septal defect (ASD) secundum closure with patch 120103. Atrial septal defect (ASD) secundum closure with patch;123463. Pacemaker system placement: permanent epicardial
>
> 120110. Sinus venosus defect (ASD) closure
>
> 120101. Atrial septal defect (ASD) secundum closure
>
> 120055. Coronary sinus interatrial communication (ASD) repair
>
> 120103. Atrial septal defect (ASD) secundum closure with patch;120102.
>
> Atrial septal defect (ASD) secundum closure with direct suture 120103. Atrial septal defect (ASD) secundum closure with patch;123246. Pericardial window creation
>
> 120143. Atrial septectomy
>
> 120103. ATRIAL SEPTAL DEFECT (ASD) SECUNDUM CLOSURE WITH PATCH
>
> 120102. Atrial septal defect (ASD) secundum closure with direct suture;121731. Aortopexy
>
> 120822. Subaortic obstruction relief
>
> 120713. Left ventricular outflow tract obstruction relief;124232. Implantable cardioverter & defibrillator (ICD) implantation: epicardial
>
> 120641. Right ventricular outflow tract obstruction relief
>
> …
>
> …
>
> ==============================
>
> Partition 2:
>
> 121640. Supravalvar aortic stenosis repair
>
> 120131. COR TRIATRIATUM (DIVIDED LEFT ATRIUM) REPAIR; 120103. ATRIAL SEPTAL DEFECT (ASD) SECUN-DUM CLOSURE WITH PATCH
>
> 120816. Closure of multiple ventricular septal defect (VSD)s 121640. Supravalvar aortic stenosis repair;121302. Pulmonary valvotomy: open
>
> 121602. Aortic valvotomy: open;121640. Supravalvar aortic stenosis repair
>
> 122911. Rastelli procedure: intraventricular left ventricle to aorta tunnel & right ventricle to pulmonary artery conduit;120803. Ventricular septal defect (VSD) closure using patch;120102. Atrial septal defect (ASD) secundum closure with direct suture
>
> 121640. Supravalvar aortic stenosis repair;121830. Aortic arch repair
>
> 120131. Cor triatriatum (divided left atrium) repair
>
> 120131. COR TRIATRIATUM (DIVIDED LEFT ATRIUM) REPAIR
>
> 120300. Mitral valvar procedure
>
> 121630. Ross procedure: aortic valve or root replacement with pulmonary autograft & pulmonary valvar replacement
>
> 120822. Subaortic obstruction relief
>
> …
>
> …

### Appendix A.2. LLM Response

Below is the full text of OpenAI’s `gpt-4-turbo` response to the above prompt:

> Partition 1 and Partition 2 present two different clustering methodologies based on a range of complex cardiopulmonary bypass (CPB) surgeries. Here are some observations that highlight differences in the clustering approach and organization of these partitions:
>
> 1. **Partition Focus**: - **Partition 1** appears to group data based on specific types and complexities of surgeries, with more focus on nuanced relationships between different surgeries common in pediatric cardiology (e.g., combinations of ASD repair methods, valve repairs, and VSD closures). - **Partition 2** appears more segmented, potentially dividing patients by broader surgical categories, like major surgery types (Aortic arch repairs, Tetralogy of Fallot repairs, Norwood procedures, etc.). This suggests a more high-level clustering compared to the specific procedural focus in Partition 1.
> 2. **Detail of Procedure Combination**: - **Partition 1** shows detailed combinations of surgeries, indicating a clustering based on specific multi-procedural patient cases (e.g., ASD closure combined with VSD closure and pulmonary artery reconstructions). - **Partition 2** while still detailed, often seems to place cases in broader procedural categories rather than intricate combinations, hinting at clustering by primary or most substantial procedure types.
> 3. **Repetition and Organization**: - **Partition 1** presents a form of clustering where similar procedures are often grouped together irrespective of the additional minor procedures, likely showing clusters based on primary intervention. - **Partition 2** regularly reuses certain procedures across different clusters, possibly categorizing based on a combination of the surgical procedure and some other clinical factors like patient demographics or surgical outcomes.
> 4. **Consistency and Case Sensitivity**: - Both partitions notably mix cases with varying capitalizations and details in the procedure descriptions, which suggests that the clustering isn’t strictly dependent on textual exactness but rather on the procedural content. However, **Partition 1** seems to tolerate more variations within clusters, while **Partition 2** may be using a stricter or different set of criteria likely incorporating additional clinical variables beyond just procedural similarity.
> 5. **Cluster Titles and Patient Outcomes**: - Neither partition explicitly titles their clusters, which requires interpretation based on the listed procedures alone. However, the clustering logic in **Partition 2** may hint at broader outcome-based or pathology-based groupings (such as focusing on all patients undergoing arterial switch operations, regardless of accompanying procedures).
> 6. **Granularity**: - **Partition 1** tends to have finer granularity, focusing on the specific procedural nuances. **Partition 2**, while detailed, appears geared towards broader surgical categories, potentially suited for analyses like resource allocation or broad outcome studies.
>
> The differences suggest diverse utility: **Partition 1** is more suitable for studies needing detailed procedural interconnections (useful in operational research, detailed post-operative complications studies) while **Partition 2** could better serve broad-brush analyses such as policy-making, general clinical outcomes, or resource planning. The choice between partitions would depend on the specific needs of the research or administrative tasks at hand.

## Appendix B. AI Explaining AI - Part 2

In this section, we present the details of the interaction with OpenAI and Google LLMs to solicit cluster labels, based on the descriptions of diagnosis and operation codes associated with each CPB. We take advantage of the ‘structured output’ feature that has been added to OpenAI’s most advanced models. For Google, we use the JSON output option that is also available for the most recent models.

### Appendix B.1. Using OpenAI’s Structured Output

OpenAI’s structured output feature is an evolution of their JSON mode. As of this writing, it is available for `gpt-4o-mini-2024-07-18` or later, or for `gpt-4o-2024-08-06` or later. See here for details.

To use this feature, we define the classes that capture what we expect to get back for each cluster. Here, we indicate to the LLM that we want to get 1) the group number (consistent with the group numbers provided in the prompt body), 2) short description of each group, 3) long description of each group. Optionally, one can add specific character/word limits for the short and long descriptions in the prompt instructions (see Appendix B.3), or otherwise modify the template as necessary for each project:

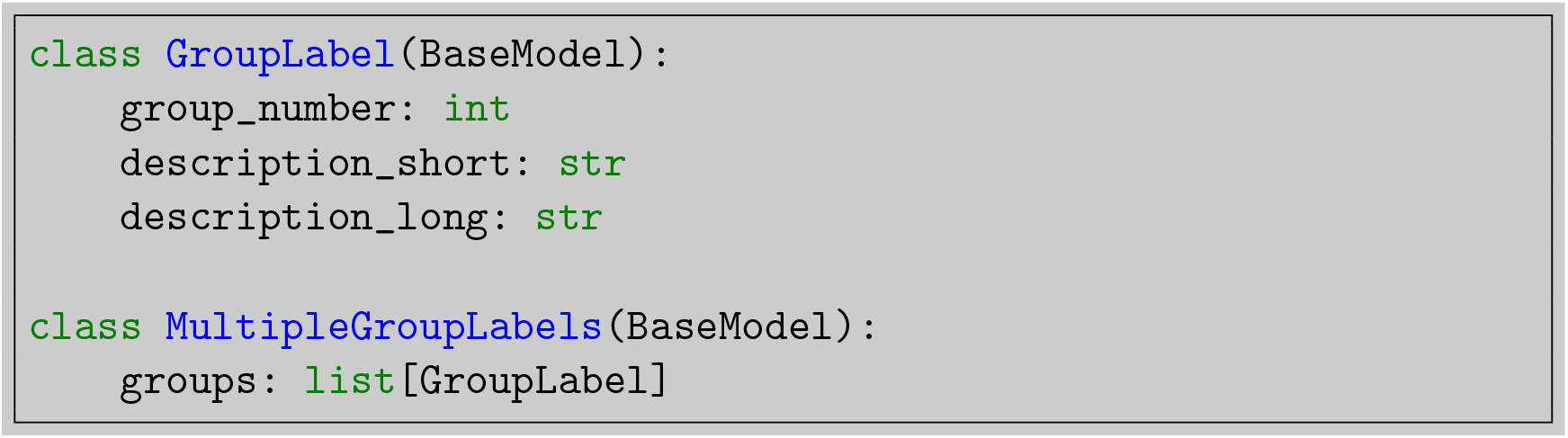

We can now submit a completion request to OpenAI by passing in the class `MultipleGroupLabels`:

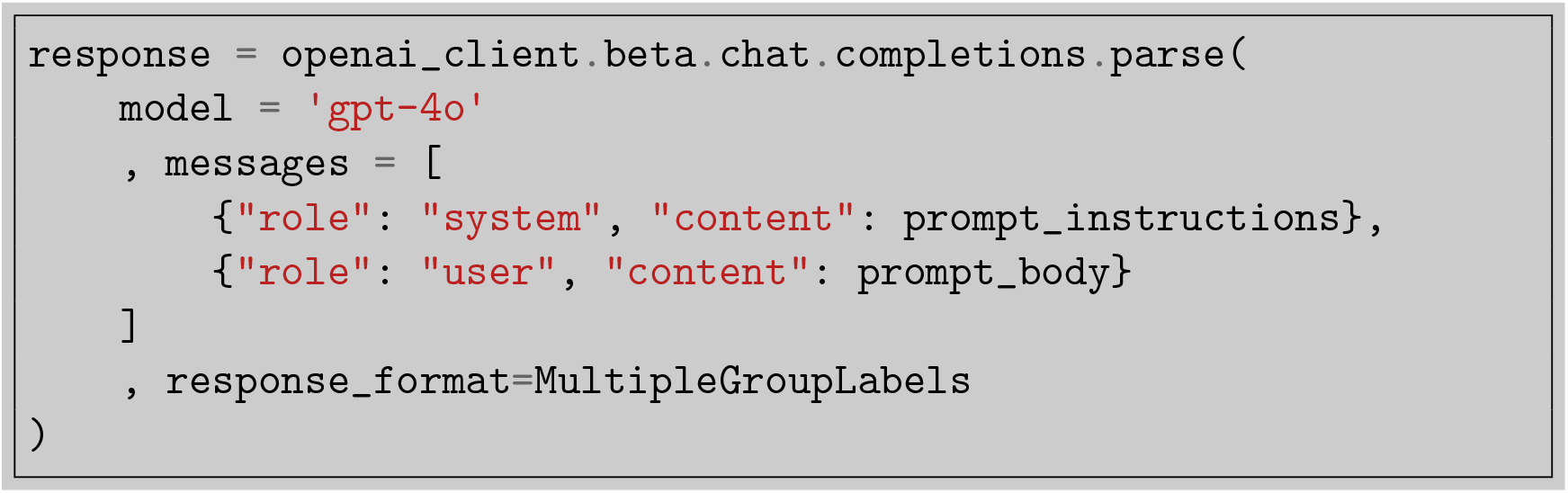

The arguments `prompt instructions` and `prompt body` are discussed further down.

### Appendix B.2. Using Google’s Structured Output

For google, we use a JSON template to convey our expectations about the response structure. Below is the schema definition:

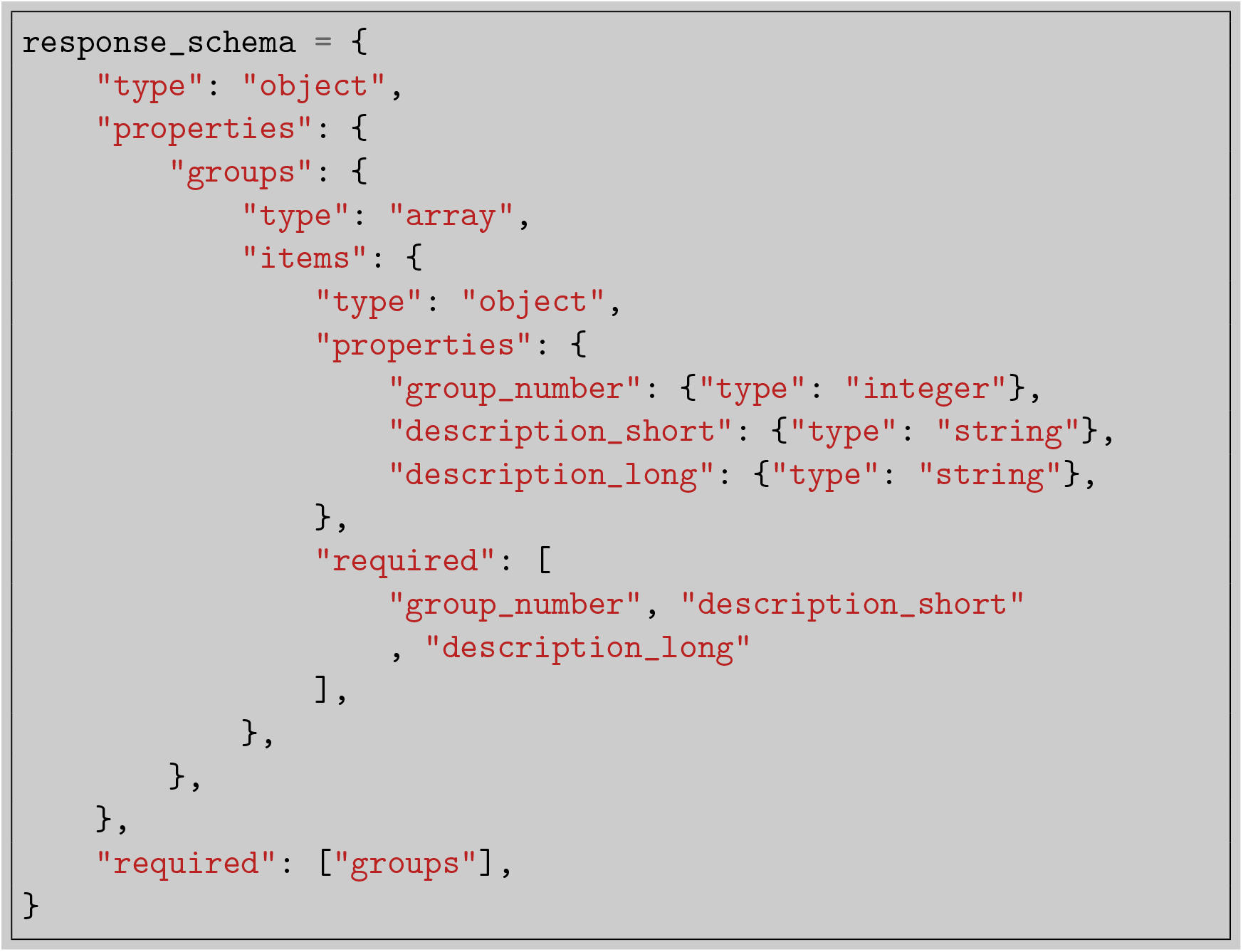

which is included in the following request:

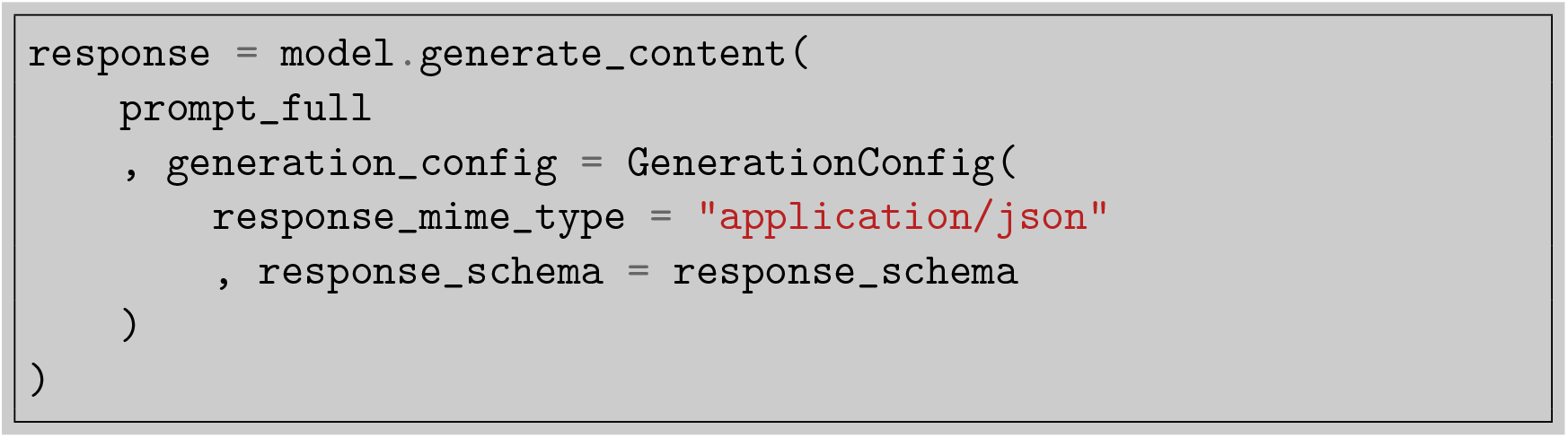

In the above, `model` is a properly instantiated `GenerativeModel` model defined in the `vertexai.generative models` module, and `prompt full` is the concatenation of `prompt instructions` and `prompt body`, discussed next.

### Appendix B.3. Prompt

The prompt consists of two sections: 1) the preamble or instructions to the LLM, and 2) the body, containing data in numbered clusters that must be described by the LLM. Below is the instructions paragraph, corresponding to the case where clusters have been created using both `diagnosis` and `operation` fields:

#### Prompt Instructions

The following is a list of 963 pediatric patients undergoing cardiopulmonary bypass. Each row contains one or more surgical procedures, separated by ’;’. These are followed by one or more diagnoses, also separated by ’;’. Patients have been grouped into 10 groups, according to their diagnoses and procedures. Please suggest group labels that are representative of their members, and also distinct from each other:

The body consists of group (or cluster) number, followed by observations in that group. For each observation, the value of the relevant text field(s) is are printed. In cases where we combine multiple text fields, we preface each text field with the name ofthe field. Below is an excerpt to illustrate the point. (*Italics* and **boldface** are added for highlighting in the manuscript, and prompts submitted to LLMs are plain text.)

#### Prompt Body

**Group 1:**

*Operations*: aortic root replacement: valve sparing technique || *Diagnoses*: aortic regurgitation; congenital anomaly of aortic valve; doubly committed juxta-arterial ventricular septal defect (vsd) with anteriorly malaligned fibrous outlet septum and perimembranous extension

*Operations*: scimitar syndrome (partially anomalous pulmonary venous connection) repair || *Diagnoses*: partial anomalous pulmonary venous connection of scimitar type

*Operations*: aortic valvar replacement using mechanical prosthesis; left ventricular outflow tract obstruction relief || *Diagnoses*: aortic regurgitation; lv outflow tract obstruction; aortic valvar stenosis - congenital; discordant va connections (tga); superior caval vein (svc) abnormality

*Operations*: mitral valvar replacement; mitral valvar annuloplasty||

*Diagnoses*: mitral regurgitation

*Operations*: mitral valvar procedure || *Diagnoses*: mitral regurgitation; mitral valvar abnormality; mitral valvar prolapse

…

…

=======

**Group 2:**

*Operations*: vsd closure; patent arterial duct (pda) closure: surgical ||*Diagnoses*: perimembranous central ventricular septal defect (vsd); patent arterial duct (pda)

…

…

## Appendix C. Cluster Mapping Algorithm

**Figure C.1.**
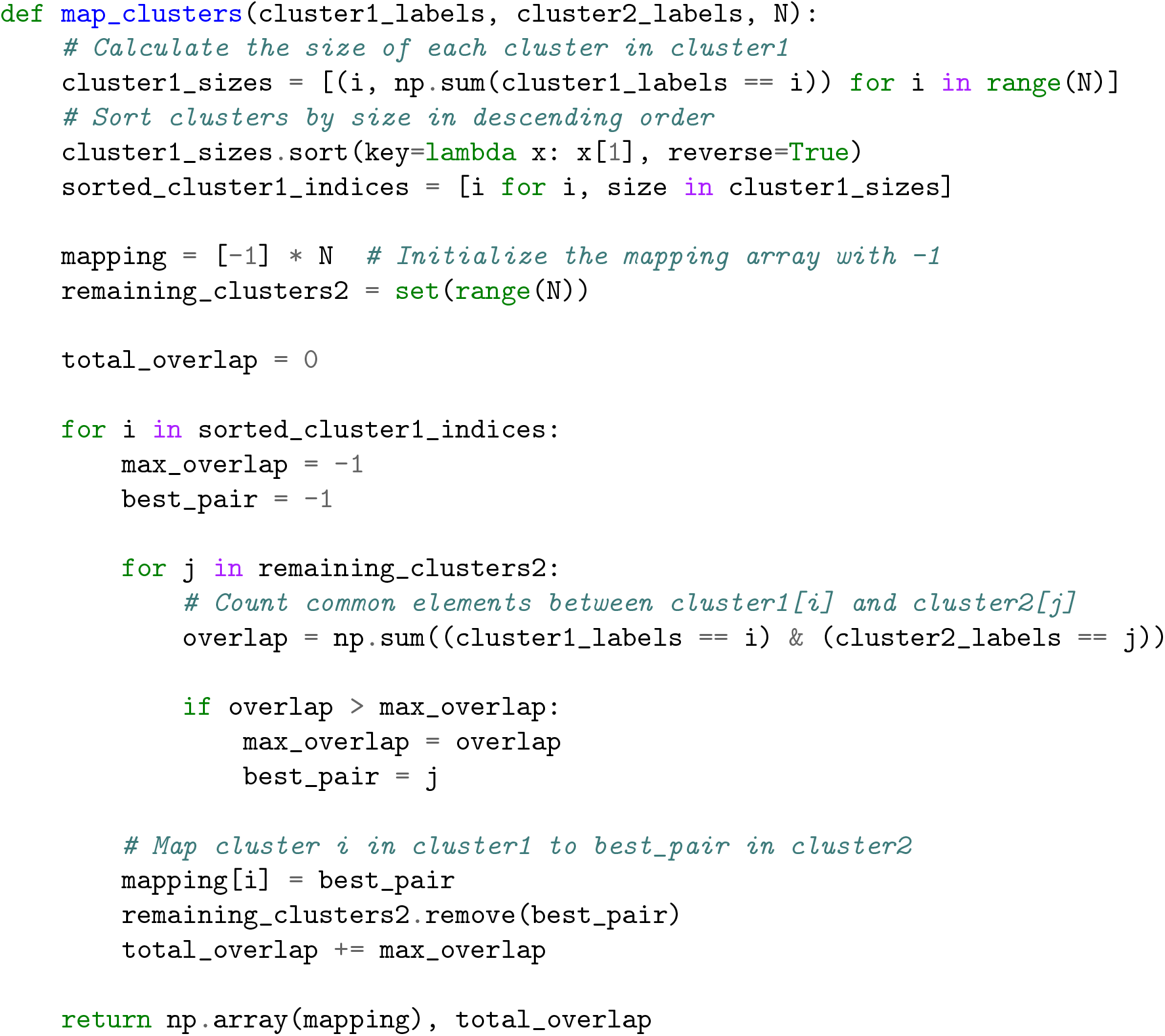
Python implementation of an algorithm - created by the authors - for mapping two data partitions to maximise total member overlap. The algorithm is ‘greedy’ since it proceeds down a sorted list of clusters in one partition to achieve an *O*(*N* ^2^) scaling behaviour, instead of *O*(*N* !), where *N* is the number of clusters in each partition.

Figure C.1 lists the Python implementation of our algorithm for mapping clusters between two different clustering results. It uses a greedy, *O*(*N* ^2^), approach for maximising the overlap between corresponding clusters in the two sets. An exhaustive search of all possible mappings between two clusters would be an *O*(*N* !) solution. Instead, our algorithm sorts one cluster set in descending size, and begins matching the second cluster set to the first one by one, in each step searching among the remaining clusters of the second set to find one that has maximum overlap with the cluster of focus in the first set.

This algorithm is particularly useful in evaluating the stability of clustering algorithms or comparing the results of different clustering methods. By mapping clusters based on their overlap, it provides a quantitative way to assess how similar two clustering results are, which is essential in fields like data mining, bioinformatics, and pattern recognition.

## Appendix D. Cluster Mapping Full Results

The table containing the full set of results for mapping of cluster labels across ten clustering runs has been provided as *cluster mapping full results*.*csv*. It is similar to the table in Table 4, but it has 10 columns of cluster labels, five produced by OpenAI’s `gpt-4o` model (columns ending in even digits) and five produced by Google’s `gemini-1.5-pro` model (columns ending in odd digits).

